# Choroid plexus asymmetry on routine MRI as an imaging signature of breast cancer brain metastasis

**DOI:** 10.64898/2026.10.13.26365386

**Authors:** Daiying Lin, Tianxiang Zhu, Xiangyuan Ma, Yiru Hu, Yurui Hao, Zonghuan Cai, Shaoxian Chen, Danfeng Wang, Mu Li, Xiaofeng Chen, Jianhui Li, Yulong Lin, Haoyang Huang, Zhiqi Yang, Gang Xiao, Zhuozhi Dai

## Abstract

**Background:** Brain metastasis affects 15–30% of patients with breast cancer, yet brain-specific biology underlying central nervous system colonisation is poorly captured by available clinical measures. The choroid plexus, an MRI biomarker of neuroinflammation and a primary immune-cell gateway implicated in brain metastasis, has not been examined in this setting.

**Methods:** In a real-world, cross-sectional study of 1,000 women with breast cancer (190 with brain metastasis), bilateral choroid plexus volumes on contrast-enhanced brain MRI were quantified using a deep-learning pipeline. Guided by the principle that hemispheric asymmetry indices are sensitive to focal pathology, this study derived from an exploratory bilateral volume comparison a parameter-free Choroid Plexus Asymmetry Index (CPAI = |L−R|/(L+R)). Multivariable logistic regression, propensity-score matching, laterality and grade-stratified analyses assessed robustness.

**Results:** The results show that total choroid plexus volume does not differ between groups, whereas left and right volumes change asymmetrically and CPAI is elevated in patients with metastasis (median 0.088 vs 0.059; Cliff’s δ = 0.20, 95% CI 0.10–0.29). CPAI is minimally correlated with 10 peripheral markers and with histological grade. The association is robust under multivariable adjustment (adjusted OR 1.58 per +1 SD, 95% CI 1.32–1.88) and propensity-score matching, with |CPAI| scaling with metastatic burden, independent of metastasis side.

**Conclusions:** Choroid plexus morphological asymmetry on routine brain MRI is associated with breast cancer brain metastasis independently of peripheral inflammation, tumour grade and conventional markers.

## Introduction

Brain metastasis (BM) is among the most devastating complications of breast cancer, with 15–30% of patients developing BM during their disease course [1,2] and median post-diagnosis survival of 4–16 months [3]. Although systemic inflammatory and tumour markers offer some prognostic guidance, the brain-specific biological processes that accompany metastatic involvement of the central nervous system (CNS) remain poorly captured by available clinical measures [4,5].

Once viewed simply as the site of cerebrospinal fluid (CSF) production, the choroid plexus (CP) has emerged as a dynamic immune command centre and a primary CNS immune-cell entry site [6,7], with a blood–CSF barrier molecularly distinct from the blood–brain barrier and potentially vulnerable to cancer-related perturbations [8,9]. Converging mechanistic evidence implicates the CP in CNS metastatic spread through barrier disruption, trans-barrier migration, ciliary damage, neurotransmitter reprogramming and vascular remodelling at the blood–CSF interface [10–14]. Although CP imaging is an established quantitative neuroinflammation biomarker across a range of neuropsychiatric conditions [15,16], no human imaging study has tested whether the CP alterations documented in preclinical brain-metastasis models produce detectable signatures in patients.

Peripheral markers of systemic inflammation and oxidative stress are prognostic in breast cancer: the neutrophil-to-lymphocyte ratio (NLR) and monocyte-to-lymphocyte ratio (MLR) are associated with outcomes [17], and HDL-C–based ratios (MHR, NHR, LHR) serve as composite inflammation– oxidative-stress markers [18,19]. Whether these circulating markers capture brain-specific metastatic processes, or instead index a parallel systemic axis, remains uncertain, motivating a CP-based imaging readout that interrogates the metastatic gateway directly.

Choosing such a readout is not arbitrary. When a pathological process is focal or spatially heterogeneous, hemispheric asymmetry indices (|L−R|/(L+R)) are more sensitive disease readouts than mean volumes, a principle exemplified by hippocampal subfield asymmetry in Alzheimer’s disease [20] and reinforced by whole-brain [21] and ENIGMA Laterality Working Group analyses [22]. These considerations identify hemispheric asymmetry as a plausible readout for the CP, whose cellular composition is intrinsically heterogeneous [6] and whose perturbation in cancer metastasis is mechanistically focal [11,12].

This article presents a discovery-phase imaging study of CP morphology in breast cancer patients with versus without brain metastasis, using routine contrast-enhanced brain MRI and an automated deep-learning segmentation pipeline. An exploratory bilateral volume comparison revealed a hemisphere-specific, not global, pattern, which was formalised as the parameter-free Choroid Plexus Asymmetry Index (CPAI = |L − R|/(L+R)). Two pre-specified evaluations were then conducted: (1) statistical robustness of the CPAI signal under confounder adjustment; and (2) its relationship to circulating inflammation–oxidative-stress information, specifically whether it carries non-redundant information. Because BM was prevalent at imaging, the study is framed as discovery-phase rather than mechanistic.

## Methods

### Study Design and Participants

This real-world cross-sectional study was conducted at Shantou Central Hospital between January 2018 and April 2024, following STROBE guidelines; all data were derived from routinely acquired clinical brain MRI and laboratory records. Consecutive female breast cancer patients who underwent clinically indicated brain MRI were screened for eligibility. Inclusion criteria were: (1) histologically confirmed breast cancer; (2) contrast-enhanced brain MRI of adequate quality for choroid plexus (CP) assessment; and (3) complete blood count, lipid panel and tumour-marker data available within 30 days of imaging. Exclusion criteria were primary brain tumours, prior cranial surgery or radiation therapy, other intracranial lesions (cysts, arteriovenous malformations, infarction, contusion and laceration, abscess, demyelinating lesions), significant motion artefacts precluding reliable segmentation, incomplete documentation, and, for the control group, skull metastasis. Of 1,281 initially screened patients, 281 were excluded on these criteria, yielding 1,000 eligible patients (190 with brain metastasis [BM], 810 without).

Brain metastasis was defined as the presence of one or more intra-axial enhancing lesions on contrast-enhanced MRI, confirmed by the treating neuroradiologist and consistent with metastatic disease in the clinical context. For the laterality sensitivity analysis, BM lesions were further classified as left-only, right-only, bilateral or midline. The Institutional Review Board of Shantou Central Hospital approved the study (approval number: [2026] Research No. 051) with waiver of informed consent.

### Choroid Plexus Volumetry and Asymmetry Quantification

Contrast-enhanced three-dimensional T1-weighted brain MRI was acquired using a standardised institutional protocol (acquisition parameters in Supplementary Methods). The CP was automatically segmented in both lateral ventricles using a deep-learning pipeline developed upon the nnU-Net framework [23], retrained and fine-tuned on in-house contrast-enhanced 3D T1-weighted MRI. The optimised algorithm achieved a 3D Dice coefficient of 0.83 and HD95 (95th percentile Hausdorff distance) of 1.90 mm, significantly outperforming existing methods (all P < 0.01; Figure S1 and Table S9 in the Supplementary Material). All segmentation outputs were verified by trained neuroradiologists blinded to clinical status.

Left, right and total (left + right) CP volumes (mm^3^) were extracted as the primary morphological measurements. Exploratory bilateral volume comparison (see Results) indicated that BM-associated volumetric alterations were hemisphere-specific rather than global, motivating derivation of the Choroid Plexus Asymmetry Index: CPAI = |Left − Right| / (Left + Right), yielding values from 0 (perfect symmetry) to 1 (complete unilaterality). The index is parameter-free and was used as the morphological predictor in all subsequent inferential analyses.

### Laboratory Biomarkers

Peripheral blood samples obtained within 30 days of MRI were used to compute 10 systemic inflammation and inflammation–oxidative-stress markers (NLR, MLR, PLR, SII, SIRI, PIV, NHR, LHR, MHR and PHR) and two tumour markers (CEA and CA15-3); definitions and formulae are provided in Supplementary Methods. Monocyte counts were reported to a resolution of 0.1 × 10^9^/L; the six patients (0.6%) with a reported count of zero were assigned half that limit (0.05 × 10^9^/L) before the monocyte-derived markers (MLR, SIRI, PIV and MHR) were computed.

### Statistics and Reproducibility

#### Statistical analysis

Between-group comparisons used the Mann–Whitney U test for continuous variables and the χ^2^ test for categorical variables, with standardised mean differences reported alongside P-values. Continuous predictors were log-transformed where skewed and standardised to z-scores, so odds ratios (ORs) are interpreted per +1 SD. The primary model was a multivariable logistic regression with forced inclusion of CPAI, age, log diagnostic-to-MRI interval, tumour markers (CEA, CA15-3) and two representative peripheral markers (MLR, LHR; selected after collinearity diagnostics, all variance inflation factors < 1.50); logit-linearity was verified by restricted cubic splines. Robustness was assessed by four pre-specified analyses (parsimonious model; 1:1 optimal propensity-score matching on age and log diagnostic interval [24]; laterality analyses; histological grade stratification), and two pre-specified secondary analyses (single-marker decomposition; stratified CPAI × marker interactions) with Benjamini–Hochberg false-discovery-rate control. Analyses used Python 3.12 and R 4.2.3; two-sided P < 0.05 defined statistical significance. Full statistical protocols are provided in Supplementary Methods.

## Results

### Patient characteristics

Of 1,000 eligible breast cancer patients, 190 had documented BM and 810 did not (Table 1). Age was similar between groups (mean 55.0 ± 11.3 vs 54.3 ± 10.4 years, P = 0.560, SMD = 0.067). The diagnostic-to-MRI interval was longer in BM patients, as expected when brain MRI is performed later in the disease course of patients with metastasis; this imbalance was forced into all multivariable models in log-transformed form (see Methods). Tumour markers CEA and CA15-3 and most peripheral inflammation indices were higher in BM patients; LHR was lower.

**Table 1.** Baseline characteristics, full cohort (n = 1,000).

| Characteristic | non-BM (n = 810) | BM (n = 190) | P-value | SMD |
| --- | --- | --- | --- | --- |
| Age (years) | 55.0 ± 11.3 | 54.3 ± 10.4 | 0.560 | 0.067 |
| Diagnostic-to-MRI interval (d) | 7.6 (2.9–185.2) | 825.8 (200.2–1,454.8) | <0.001 | 0.909 |
| Left CP (mm<sup>3</sup>) | 516 (405–623) | 528 (370–621) | 0.580 | 0.078 |
| Right CP (mm<sup>3</sup>) | 511 (408–618) | 486 (366–600) | 0.021 | 0.162 |
| Total CP (mm<sup>3</sup>) | 1,034 (826–1,243) | 1,017 (741–1,194) | 0.120 | 0.125 |
| NLR | 2.45 (1.79–3.57) | 3.14 (2.06–4.82) | <0.001 | 0.339 |
| MLR | 0.21 (0.16–0.29) | 0.29 (0.20–0.44) | <0.001 | 0.517 |
| PLR | 156 (121–209) | 174 (118–262) | 0.007 | 0.143 |
| SII | 652 (428–1,002) | 771 (372–1,407) | 0.075 | 0.070 |
| SIRI | 0.90 (0.57–1.42) | 1.11 (0.67–2.31) | <0.001 | 0.303 |
| PIV | 227 (136–406) | 263 (112–662) | 0.095 | 0.108 |
| NHR | 2.96 (2.17–4.10) | 3.12 (2.07–4.98) | 0.226 | 0.081 |
| LHR | 1.16 (0.87–1.56) | 0.96 (0.70–1.42) | <0.001 | 0.333 |
| MHR | 0.26 (0.19–0.35) | 0.29 (0.21–0.41) | 0.004 | 0.216 |
| PHR | 183 (142–242) | 176 (116–244) | 0.102 | 0.181 |
| CEA | 2.50 (1.50–4.50) | 5.35 (2.62–16.02) | <0.001 | 0.646 |
| CA15-3 | 15.7 (9.9–26.0) | 28.5 (15.8–115.1) | <0.001 | 0.672 |

Continuous variables presented as median (IQR) or mean ± SD. BM, brain metastasis; NLR, neutrophil-to-lymphocyte ratio; MLR, monocyte-to-lymphocyte ratio; PLR, platelet-to-lymphocyte ratio; SII, systemic immune-inflammation index; SIRI, systemic inflammation response index; PIV, pan-immune-inflammation value; NHR, neutrophil-to-HDL-C ratio; LHR, lymphocyte-to-HDL-C ratio; MHR, monocyte-to-HDL-C ratio; PHR, platelet-to-HDL-C ratio; SMD, standardised mean difference. SMDs for the skewed blood-based biomarkers (NLR, MLR, PLR, SII, SIRI, PIV, NHR, LHR, MHR, PHR, CEA and CA15-3) were computed on log-transformed values; SMDs for age, the diagnostic-to-MRI interval and CP volumes were computed on the raw scale. Diagnostic-to-MRI interval reflects the longer clinical follow-up window characteristic of patients with established metastatic disease and was log-transformed and forced into all multivariable models.

### Hemisphere-specific CP morphology and definition of CPAI

Total, left, and right CP volumes were first compared between BM and non-BM patients (Figure 1A; Table 1). Total CP volume did not differ between groups (Cohen’s d = −0.13, P = 0.120). Left and right CP volumes showed hemisphere-specific patterns in BM patients: a small group-level difference was detected on the right side (median 486 vs 511 mm^3^; Cohen’s d = −0.17, P = 0.021), whereas the left CP did not differ (median 528 vs 516 mm^3^, P = 0.580). To formalise this hemispheric asymmetry into a single, parameter-free index, the CPAI = |L − R|/(L+R) was calculated for each subject. CPAI was elevated in BM patients (median 0.088 vs 0.059; Cliff’s δ = 0.20 [95% CI 0.10–0.29], P = 2.7 × 10^−5^; Figure 1B; an illustrative high-CPAI case is shown in Figure 1C).

**Figure 1.**
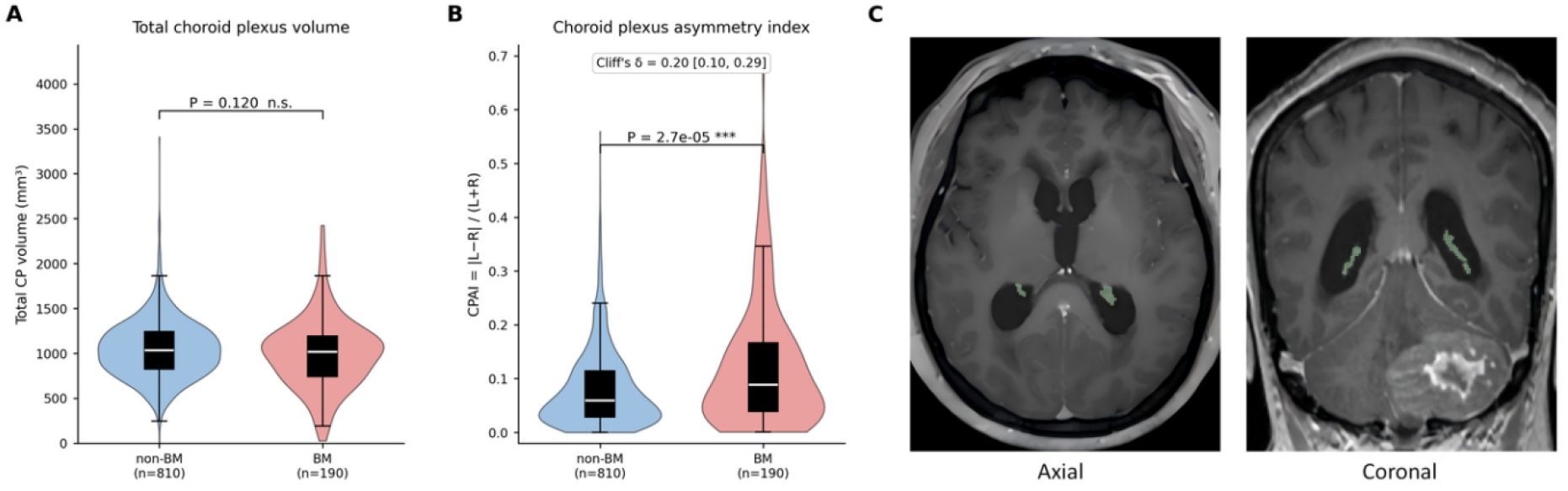
Choroid plexus volume, choroid plexus asymmetry index (CPAI) distribution, and an illustrative high-CPAI case. **(A)** Violin/box plots comparing total CP volume between non-BM (n = 810) and BM (n = 190) groups. Total CP volume did not differ between groups (Cohen’s d = −0.13, P = 0.120). **(B)** Violin/box plots comparing CPAI distribution between non-BM and BM groups. BM patients showed significantly higher CPAI (Cliff’s δ = 0.20 [95% CI 0.10–0.29]; P = 2.7 × 10^−5^). **(C)** Representative axial (left) and coronal (right) contrast-enhanced T1-weighted images from a BM patient with high CPAI (illustrative case, CPAI = 0.672). Automated CP segmentation (green overlay) shows a visibly enlarged left CP relative to the right, directly illustrating the hemispheric divergence quantified by CPAI = |L−R|/(L+R).

### Univariate associations and cross-correlation structure

CPAI was strongly associated with BM in univariate logistic regression (OR = 1.53 per +1 SD, 95% CI 1.33–1.77, P = 5.0 × 10^−9^; Figure 2A; Table S1). Pearson cross-correlations between CPAI and each of the 10 peripheral inflammation–oxidative-stress markers were uniformly very small (all |r| < 0.05; Figure 2B; Table S2), indicating that the imaging signal does not track peripheral information at the bivariate level. The peripheral markers, by contrast, were highly collinear among themselves (multiple pairs with |r| > 0.70; maxima SIRI – PIV r = 0.89 and SII – NLR r = 0.88; Figure 2B), motivating inclusion of only representative markers (MLR for the inflammatory–monocyte axis, LHR for the lipid–oxidative-stress axis) in the multivariable model; all VIFs were subsequently below 1.50 (Table S3).

**Figure 2.**
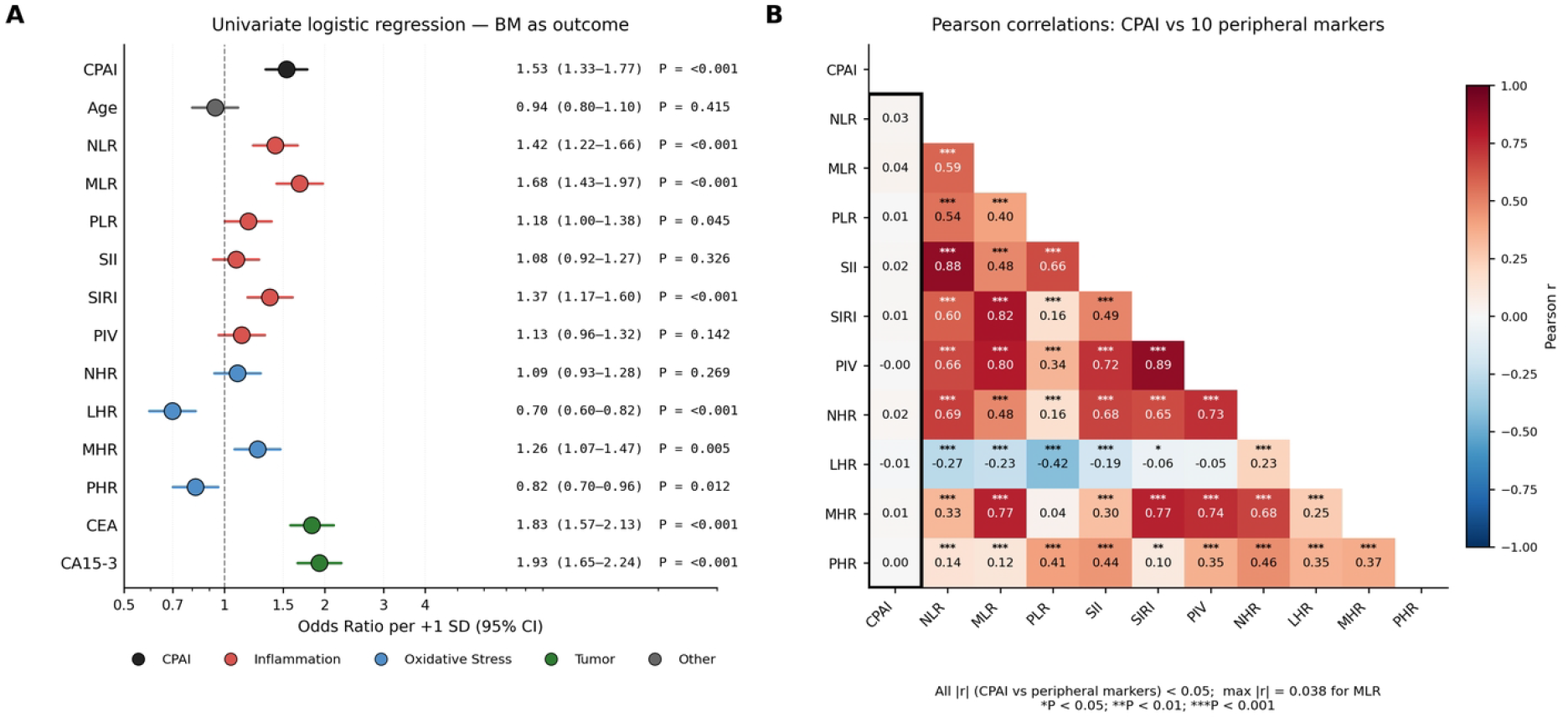
Univariate analysis and biomarker correlations (full cohort, n = 1,000). **(A)** Forest plot showing odds ratios (per +1 SD; 95% CI) for brain metastasis risk from univariate logistic regression. Biomarkers are colour-coded by category: CPAI (black), inflammation markers (red), oxidative-stress markers (blue), and tumour markers (green). CPAI demonstrated a strong univariate association (OR = 1.53, 95% CI 1.33–1.77, P = 5.0 × 10^−9^). **(B)** Pearson correlation heatmap among CPAI and the 10 systemic biomarkers. High correlations were observed among several inflammation marker pairs (e.g., SIRI–PIV r = 0.89; SII–NLR r = 0.88), justifying selection of representative markers for multivariable modelling. CPAI showed only minimal cross-correlations with peripheral markers (all |r| < 0.05; maximum |r| = 0.038 for MLR). *P < 0.05; **P < 0.01; ***P < 0.001.

### Multivariable and sensitivity analyses

In the primary multivariable model, with forced inclusion of log(diagnostic-to-MRI interval), age, tumour markers (CEA, CA15-3) and the representative markers (MLR, LHR), the CPAI association was not attenuated, and slightly strengthened, to OR = 1.58 per +1 SD (95% CI 1.32–1.88, P = 3.6 × 10^−7^; Table 2; Figure 3A). CA15-3 and CEA remained independently associated (Table 2), and the model achieved good discrimination (C-statistic (AUC) = 0.848, AIC = 716.82). Restricted cubic spline analysis showed no departure from logit-linearity for CPAI (P = 0.25), supporting the per +1-SD OR as the appropriate continuous-effect summary.

**Table 2.** Primary multivariable model — full cohort.

| Predictor | $\beta$ | SE | OR | 95% CI | P |
| --- | --- | --- | --- | --- | --- |
| CPAI (per +1 SD) | 0.458 | 0.090 | 1.58 | 1.32–1.88 | $3.6 \times 10^{-7}$ |
| log(diagnostic interval) | 1.102 | 0.112 | 3.01 | 2.42–3.75 | $1.1 \times 10^{-22}$ |
| Age | −0.054 | 0.102 | 0.95 | 0.78–1.16 | 0.593 |
| CA15-3 | 0.415 | 0.105 | 1.51 | 1.23–1.86 | $8.1 \times 10^{-5}$ |
| CEA | 0.300 | 0.103 | 1.35 | 1.10–1.65 | 0.004 |
| MLR | 0.132 | 0.101 | 1.14 | 0.94–1.39 | 0.192 |
| LHR | −0.159 | 0.103 | 0.85 | 0.70–1.04 | 0.122 |

*C-statistic* = 0.848. *AIC* = 716.82. Continuous predictors are log-transformed where skewed and standardised to z-scores on the full cohort, so the OR is per +1 SD on the appropriate scale. CPAI, choroid plexus asymmetry index; OR, odds ratio; CI, confidence interval.

**Figure 3.**
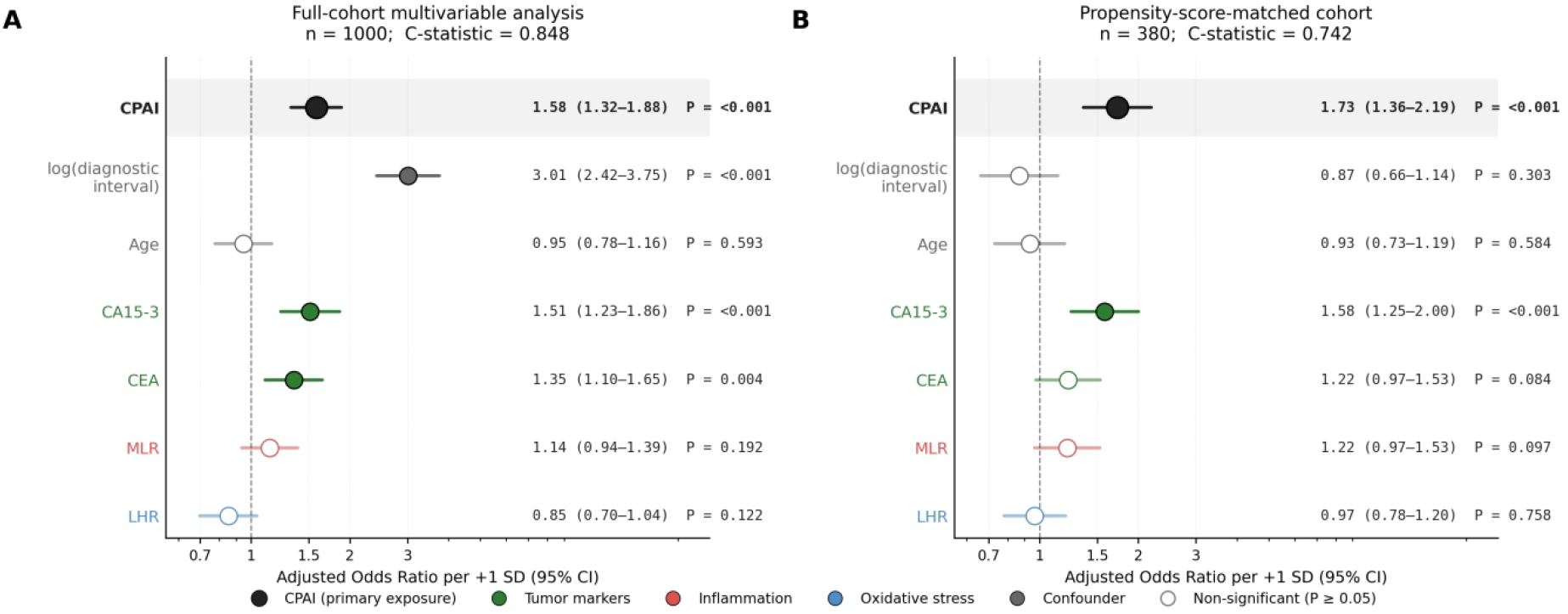
Multivariable analysis. Forest plots of adjusted odds ratios (per +1 SD; 95% CI) from multivariable logistic regression with forced inclusion of log(diagnostic-to-MRI interval), age, MLR, LHR, CEA, and CA15-3. (A) Full-cohort primary analysis: adjusted CPAI OR = 1.58 per +1 SD (95% CI 1.32–1.88; P = 3.6 × 10^−7^). (B) Propensity-score-matched sensitivity analysis: adjusted CPAI OR = 1.73 per +1 SD (95% CI 1.36–2.19; P = 7.8 × 10^−6^). The convergence between (A) and (B) supports robustness of the CPAI–BM association. *P < 0.05; **P < 0.01; ***P < 0.001.

Sensitivity analyses confirmed the robustness of the primary estimate (Figure 3; Table S4). A parsimonious model omitting the inflammation–oxidative-stress markers gave an essentially identical CPAI OR of 1.60 (95% CI 1.34–1.90, P = 1.4 × 10^−7^), and re-estimation in a propensity-score-matched cohort (n = 380, 190 matched pairs on age and log diagnostic interval) reproduced the primary estimate (multivariable OR = 1.73 per +1 SD, 95% CI 1.36–2.19, P = 7.8 × 10^−6^; Figure 3B). The convergence of these estimates across substantially different analytic strategies indicates that the CPAI–BM association is not an artefact of any particular adjustment scheme.

### Brain metastasis laterality and choroid plexus asymmetry

To verify that elevated CPAI in BM patients was not driven by a localised adjacency effect of the tumour on the ipsilateral CP, CP changes were examined by BM laterality (Figure 4; Table S5). Among patients with unilateral BM (n = 39), paired analysis revealed no significant ipsilateral CP enlargement (mean ipsilateral–contralateral difference −8.6 mm^3^; paired t P = 0.66; Wilcoxon P = 0.62; Figure 4A), and the proportion with ipsilateral predominance (48.7%) was indistinguishable from chance (binomial P = 1.00). Signed CPAI did not differ from zero in any BM subgroup after Benjamini– Hochberg correction (all q ≥ 0.20) nor across the four location subgroups (Kruskal–Wallis P = 0.11; Figure 4B), and did not predict right-only versus left-only metastasis (P = 0.28). In contrast, the magnitude of asymmetry tracked metastatic burden rather than tumour side: bilateral BM showed significantly elevated |CPAI| versus controls (median 0.103 vs 0.059, P < 0.001; Figure 4C), whereas unilateral BM did not (pooled P = 0.52; left-only P = 0.94, right-only P = 0.23). These results argue against a focal tumour-adjacency artefact and indicate that the CPAI signal scales with overall metastatic involvement.

**Figure 4.**
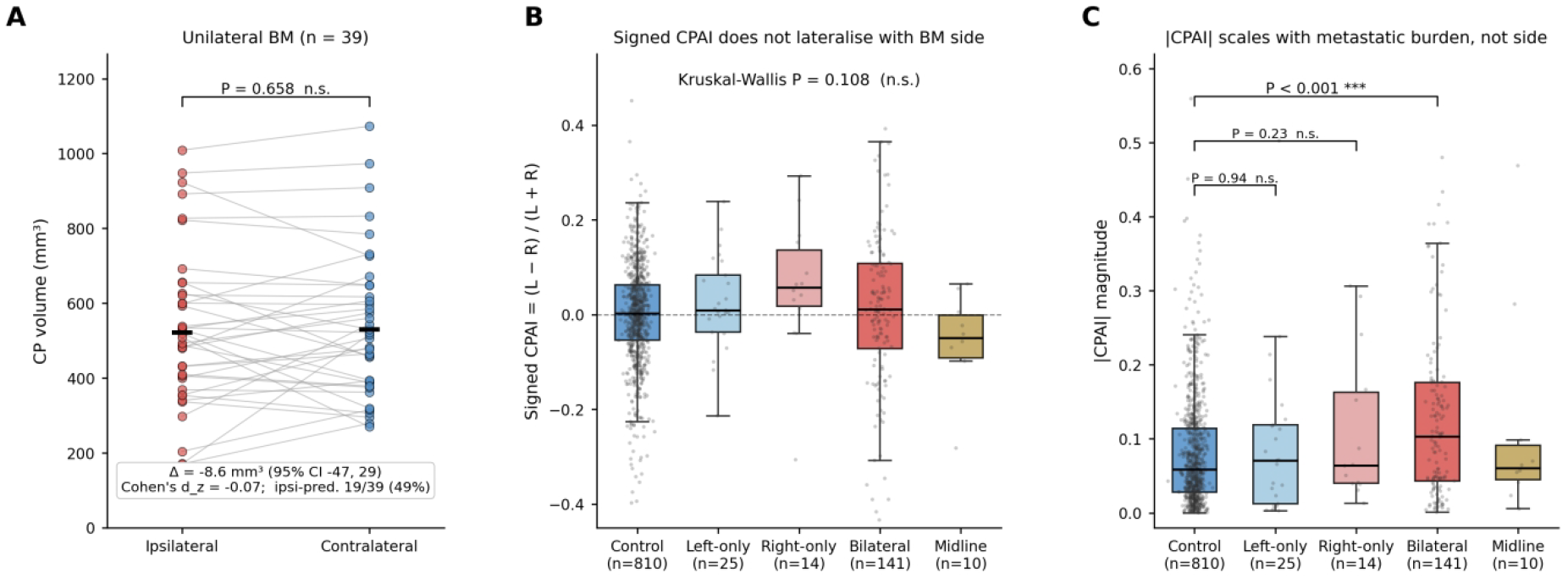
Choroid plexus asymmetry by brain metastasis laterality. **(A)** Paired comparison of ipsilateral versus contralateral CP volumes in patients with unilateral BM (n = 39). The mean ipsilateral–contralateral difference was small and not significant (paired t P = 0.66). **(B)** Box plots of signed CPAI [(L−R)/(L+R)] across control and BM subgroups; no BM subgroup differs from zero after Benjamini–Hochberg correction (all q ≥ 0.20), and the four BM subgroups do not differ from each other (Kruskal–Wallis P = 0.11). **(C)** |CPAI| magnitude by BM laterality: bilateral BM has significantly higher |CPAI| than controls (P < 0.001), whereas unilateral BM does not (pooled P = 0.52; reported in the main text and Table S5). Brackets in panel C denote the per-side comparisons versus control (left-only P = 0.94; right-only P = 0.23). Panels A–C together argue against a focal tumour-adjacency confound. NS, not significant; ***P < 0.001.

### Histological grade stratification analysis

A complementary concern, that CPAI might track primary-tumour aggressiveness rather than a CNS-compartmentalised process, was addressed by grade-stratified analysis (n = 671 with available grade, 67.1%; Table S6). CPAI was uncorrelated with grade (r = 0.004, P = 0.93); grade was itself independently associated with BM after adjusting for CPAI and CA15-3 (per-grade OR = 1.84, 95% CI 1.18 – 2.85, P = 0.007), yet adding grade to the primary model left the CPAI estimate unchanged (adjusted OR = 1.63, 95% CI 1.31–2.03, P = 9.2 × 10^−6^); and CPAI remained associated with BM in both Grade II (OR = 1.46, 95% CI 1.19–1.79, P < 0.001) and Grade III (OR = 1.91, 95% CI 1.23–2.97, P = 0.004) strata, with no effect modification (P-interaction = 0.33).

### Single-marker decomposition and CPAI × marker interaction tests

Two pre-specified secondary analyses asked whether the CPAI–BM association is concentrated in any single peripheral marker (single-marker decomposition; Table S7) or modified by any marker (stratified CPAI × marker interactions; Table S8), with multiplicity controlled by Benjamini–Hochberg FDR (q < 0.05) over the 10 markers in each analysis. Single-marker decomposition (5,000 BCa bootstrap intervals) yielded shares all below ~8% (largest: MLR 7.2%, NLR 4.6%, SIRI 4.4%), with no marker surviving FDR adjustment under either Sobel or BCa bootstrap criteria. Interaction analyses similarly yielded no significant effect modification after BH adjustment (smallest q = 0.119 for CPAI × LHR). These analyses provide no statistical evidence that CPAI’s association with BM is concentrated in, or modified by, any individual peripheral marker.

## Discussion

In 1,000 women with breast cancer, the choroid plexus shows a detectable morphological change in those with brain metastasis. The change is asymmetric, not global: total CP volume is unchanged, but CPAI is elevated in patients with metastasis. Across the two pre-specified evaluations framed in the Introduction, the CPAI signal is statistically robust under multivariable adjustment and propensity-score matching and biologically distinct from peripheral inflammation–oxidative-stress information. Whether it reflects a CNS-compartmentalised process or tracks established metastatic burden cannot be resolved cross-sectionally; the findings are discovery-phase. CPAI is not proposed as a diagnostic marker for established brain metastasis, which is directly visible on MRI; its value lies in a brain-specific biological readout and, prospectively, a candidate tool for risk anticipation in MRI-negative patients.

That an asymmetric CP signal exists at the imaging level is consistent with recent CP biology. The CP is an actively perturbable epithelial–vascular–immune interface that metastasising cancer cells exploit [25,26]: cancer-cell-derived complement C3 disrupts the blood–CSF barrier via epithelial C3aR [10]; tumour ICAM2 engages CP-epithelial ICAM1 to enable trans-barrier migration in triple-negative breast cancer [11]; CP mast cells damage epithelial cilia via tryptase [12]; tumour IL-13 drives CP GABA up-regulation, favouring leptomeningeal dissemination [13]; and leptomeningeal-metastatic cells remodel CP vasculature via extracellular vesicles [14]. Single-cell and spatial transcriptomic atlases show that the CP is spatially heterogeneous [6]. Each implicated mechanism is thus a discrete focal event on a regionally heterogeneous substrate, whose expected morphological footprint is left–right volumetric divergence rather than global change — precisely the pattern observed in this study.

This focal-perturbation logic also clarifies why an unsigned asymmetry index, rather than total CP volume or a signed left–right difference, should be the sensitive readout: focal pathology elevates the dispersion of (L−R) without consistently shifting its mean, a principle established in neuroimaging [20–22]. The CP itself sits in a ventricular system that is modestly but reliably asymmetric in healthy adults [27], establishing a small but non-zero CPAI baseline (median 0.059 in non-BM patients) on which focal pathology produces a disproportionate left–right divergence.

The focal-perturbation model also predicts that asymmetry should not track tumour side, unlike a local-adjacency hypothesis in which peritumoural mass effect [28,29] would preferentially compress the ipsilateral CP. The laterality analysis (Figure 4) adjudicates between these predictions: ipsilateral and contralateral CP volumes did not differ, ipsilateral predominance was at chance level, and signed CPAI did not predict metastasis side, whereas |CPAI| scaled with bilateral burden. This independence from lesion side parallels stroke survivors, in whom CP volume is enlarged bilaterally with no ipsilesional– contralesional difference [30], and non-lateralised CP change has been reported in multiple sclerosis [15,16,31] and neuropsychiatric lupus [32].

Turning to the first pre-specified evaluation, the CPAI–BM association is statistically robust. It was not attenuated by multivariable adjustment; restricted cubic spline analysis confirmed logit-linearity; and point estimates were stable across the full-cohort primary model, a parsimonious model and a propensity-score-matched cohort. The convergence of these analyses across substantially different adjustment strategies indicates that the association is not an artefact of any particular modelling choice.

The second pre-specified evaluation asks whether this CP signal merely tracks systemic inflammation already captured by routine blood markers. Three independent analytic axes argue against that interpretation: pairwise cross-correlations between CPAI and each of the 10 peripheral markers are uniformly small; the CPAI–BM OR is not attenuated when those markers enter the multivariable model; and single-marker decomposition and CPAI × marker interaction analyses showed that no individual marker accounted for a significant share of, or modified, the association under formal Benjamini– Hochberg multiplicity control (Tables S7, S8). This non-redundancy is biologically coherent: the blood–CSF barrier has a distinct molecular architecture from the blood–brain barrier [8], and the CP can develop a distinct intra-compartmental cytokine signature in the absence of detectable immune-cell infiltrates [33]. CPAI is likewise not a surrogate of primary-tumour aggressiveness: it was uncorrelated with histological grade, its estimate was unchanged when grade entered the model, and the association was preserved across grade strata with no interaction (Table S6).

Several practical features would facilitate the longitudinal and mechanistic studies these results motivate. CPAI is computed from routine clinical 3D T1 sequences without any additional acquisition, paralleling CP volumetric assessments already applied in Alzheimer’s disease and COVID-19 cohorts [34,35]. The deep-learning segmentation pipeline achieves accuracy comparable to recent dedicated CP networks [36], removing what has been a major reproducibility barrier in CP imaging.

This study has several limitations. First, the cross-sectional design with prevalent BM cannot establish whether CPAI predicts incident BM, the natural target of prospective longitudinal studies; reverse causation cannot be excluded, although the low cross-correlations with peripheral markers indicate that CPAI captures CNS-compartmentalised information not reducible to systemic inflammation. Second, all patients came from a single institution, so external validation is essential. Third, although parameter-free, the CPAI formula was crystallised from an exploratory hemispheric comparison within this cohort; independent replication as an a priori predictor is essential. Finally, histological grade was unavailable for 32.9% of patients and the unilateral-BM subgroup was small (n = 39). Prospective longitudinal cohorts measuring CPAI before incident BM, together with CSF biomarkers and functional CP imaging (arterial spin labelling, TSPO-PET), would extend CPAI from imaging signature toward mechanistic interrogation [14].

In summary, this discovery-phase imaging study identifies CP morphological asymmetry as an imaging signature of breast cancer brain metastasis. The signal is hemisphere-specific rather than global, and it is statistically distinct from circulating inflammation–oxidative-stress information. Because CPAI is derivable from routine clinical MRI using automated segmentation, it is well suited for prospective longitudinal validation in independent cohorts, opening a line of inquiry into the choroid plexus as both a quantitative window on CNS-compartmentalised involvement and a potential mechanistic node in the host response to brain metastasis.

## Data Availability

All data produced in the present study are available upon reasonable request to the authors.

## Data Availability

The de-identified imaging-derived measurements (left CP volume, right CP volume, total CP volume, CPAI) and laboratory variables that support the findings of this study are available from the corresponding author upon reasonable request, subject to approval by the Institutional Review Board and to the restrictions imposed by patient-privacy regulations governing real-world clinical data. The raw brain MRI scans cannot be shared publicly because they constitute protected health information under the local data-protection framework. Source data underlying Figs. 1 – 4 are available from the corresponding author upon reasonable request.

## Code Availability

The deep-learning segmentation pipeline was built on the open-source nnU-Net framework (https://github.com/MIC-DKFZ/nnUNet). Custom analysis code (Python 3.12 and R software 4.2.3) used to compute CPAI and to generate all reported statistics, figures and tables is available from the corresponding author upon reasonable request.

## Ethics approval

This study was performed in accordance with the Declaration of Helsinki and approved by the Institutional Review Board of Shantou Central Hospital (approval number: [2026] Research No. 051; date of approval: 26 June 2026). The requirement for individual informed consent was waived by the Institutional Review Board because the study retrospectively analysed de-identified, routinely collected clinical data.

## CRediT authorship contribution statement

Daiying Lin: Methodology, Data curation, Formal analysis, Writing – original draft. Tianxiang Zhu: Methodology, Data curation, Formal analysis, Writing – original draft. Xiangyuan Ma: Methodology, Data curation, Formal analysis, Writing – original draft. Yiru Hu: Data curation, Formal analysis. Yurui Hao: Data curation, Formal analysis. Zonghuan Cai: Investigation, Resources. Shaoxian Chen: Investigation, Resources. Danfeng Wang: Investigation, Resources. Mu Li: Investigation. Xiaofeng Chen: Investigation, Resources. Jianhui Li: Investigation, Resources. Yulong Lin: Investigation, Resources. Haoyang Huang: Investigation, Resources. Zhiqi Yang: Conceptualization, Supervision, Writing – review & editing. Gang Xiao: Conceptualization, Supervision, Writing – review & editing. Zhuozhi Dai: Conceptualization, Supervision, Writing – review & editing.

## Declaration of generative AI and AI-assisted technologies in the manuscript preparation process

During the preparation of this work the authors used Kimi (Moonshot AI) in order to assist with language editing and with formatting the manuscript according to journal guidelines. After using this tool, the authors reviewed and edited the content as needed and take full responsibility for the content of the published article.

