## Supplemental Information for "Choroid plexus asymmetry on routine MRI as an imaging signature of breast cancer brain metastasis"

### Contents

**Supplementary Methods**

1. MRI acquisition
2. Laboratory biomarker definitions
3. Statistical analysis

**Supplementary Figure**

Figure S1. Visual Comparison of Choroid Plexus Segmentation Performance Across Different Deep Learning Models

**Supplementary Tables**

Table S1. Univariate logistic regression for brain metastasis

Table S2. Cross-correlation between CPAI and the ten peripheral markers

Table S3. Collinearity diagnostics for the primary multivariable model

Table S4. Sensitivity analyses of the CPAI–brain metastasis association

Table S5. Choroid plexus volume and asymmetry by brain metastasis laterality

Table S6. CPAI–brain metastasis association stratified by histological grade

Table S7. Single-marker decomposition of the CPAI–brain metastasis association

Table S8. Stratified CPAI × marker interaction analyses

Table S9. Performance Comparison of Choroid Plexus Segmentation Models Using Five-Fold Cross-Validation

### Supplementary Methods

#### 1. MRI acquisition

All examinations were performed at Shantou Central Hospital on a 3.0 T system (Siemens Healthcare, Erlangen, Germany) with an 18-channel head coil. Contrast-enhanced three-dimensional T1-weighted images, the sequence from which all choroid plexus measurements were derived, were acquired with a magnetisation-prepared rapid gradient-echo (MP-RAGE) sequence obtained 5 minutes after intravenous administration of gadopentetate dimeglumine (Magnevist, Bayer HealthCare, Berlin, Germany) at a dose of 0.1 mmol/kg.

Acquisition parameters were repetition time 1900 ms, echo time 2.48 ms, inversion time 900 ms, flip angle 9°, receiver bandwidth 170 Hz per pixel, field of view 256 × 256 mm, reconstructed voxel size 1 × 1 × 1 mm, and slice thickness 1 mm with no interslice gap, covering the whole brain in sagittal sections. Total acquisition time was about 4 min.

### 2. Laboratory biomarker definitions

| Abbreviation | Full name | Formula | Axis |
|---|---|---|---|
| NLR | Neutrophil-to-lymphocyte ratio | neutrophil / lymphocyte | Inflammation |
| MLR | Monocyte-to-lymphocyte ratio | monocyte / lymphocyte | Inflammation |
| PLR | Platelet-to-lymphocyte ratio | platelet / lymphocyte | Inflammation |
| SII | Systemic immune-inflammation index | platelet × neutrophil / lymphocyte | Inflammation |
| SIRI | Systemic inflammation response index | neutrophil × monocyte / lymphocyte | Inflammation |
| PIV | Pan-immune-inflammation value | neutrophil × platelet × monocyte / lymphocyte | Inflammation |
| NHR | Neutrophil-to-HDL-C ratio | neutrophil / HDL-C | Oxidative stress |
| LHR | Lymphocyte-to-HDL-C ratio | lymphocyte / HDL-C | Oxidative stress |
| MHR | Monocyte-to-HDL-C ratio | monocyte / HDL-C | Oxidative stress |
| PHR | Platelet-to-HDL-C ratio | platelet / HDL-C | Oxidative stress |

Counts are absolute values from the same complete blood count.

3. Statistical analysis

**General conventions.** Continuous variables are summarised as median (interquartile range) or mean ± SD. Between-group comparisons used the Mann–Whitney U test for continuous variables and the $\chi^2$ test for categorical variables; standardised mean differences (SMD) were reported alongside P-values. Continuous predictors entering regression models were log-transformed where skewed and standardised to z-scores on the full cohort, so that odds ratios (ORs) are interpreted per +1 SD. All analyses used Python 3.12 and R software 4.2.3; two-sided $P < 0.05$ defined statistical significance unless otherwise specified.

**Exploratory volume comparison.** Left, right and total CP volumes were compared between BM and non-BM groups using Mann–Whitney U tests. Effect sizes were quantified using Cohen's d for the bilateral volume comparisons (left, right, total CP). For the CPAI comparison, Cliff's $\delta$ was additionally computed, with 95% confidence interval obtained from 5,000 nonparametric bootstrap resamples.

**Univariate associations and cross-correlation analysis.** Univariate logistic regressions estimated ORs per +1 SD for each predictor against BM status. To characterise the correlation structure among all candidate predictors, pairwise Pearson correlations were computed among CPAI and the 10 peripheral inflammation–oxidative-stress markers.

**Primary multivariable model.** Multivariable logistic regression was performed on the full cohort (n = 1,000) with forced inclusion of CPAI, age, log-transformed diagnostic-to-MRI interval, tumour markers (CEA, CA15-3) and two representative peripheral markers (MLR for the inflammatory–monocyte axis, LHR for the lipid–oxidative-stress axis). MLR and LHR were selected on the basis of pairwise Pearson correlations among the 10 peripheral markers: high collinearity ($|r| > 0.70$ for 9 of 45 pairs) precluded inclusion of all 10; the selection was subsequently confirmed by variance inflation factors (VIFs) all below

1.50. Model discrimination was assessed by the C-statistic (area under the ROC curve) with 95% CIs from DeLong’s method; model fit was evaluated by the Akaike information criterion (AIC). Departure from logit-linearity in the CPAI–BM association was assessed by likelihood-ratio comparison of the linear model against a restricted cubic spline model with four knots (5th, 35th, 65th and 95th percentiles of CPAI).

**Robustness and sensitivity analyses.** Four pre-specified analyses assessed the robustness of the primary CPAI–BM association. ***(i) Parsimonious model.*** A reduced model omitting the inflammation–oxidative-stress markers (CPAI + age + log diagnostic-to-MRI interval + CEA + CA15-3) tested whether the CPAI estimate depended on their inclusion. ***(ii) Propensity-score matching.*** Propensity scores were estimated by logistic regression with log-transformed diagnostic-to-MRI interval and age as covariates [24]; 1:1 optimal matching without replacement, globally minimising the total within-pair distance in logit propensity score, was performed using a caliper of 0.25 SD of the logit propensity score; all 190 BM patients were matched. Balance was verified by SMD < 0.10; sensitivity to caliper width (0.15–0.30) was confirmed. To facilitate comparison, PSM-cohort ORs are reported per +1 full-cohort SD (full-cohort CPAI SD ≈ 0.084; PSM-cohort SD ≈ 0.098 owing to BM enrichment). ***(iii) Laterality analysis.*** To test whether CPAI elevation could be attributable to a localised tumour-adjacency effect on the ipsilateral CP, the following analyses were performed. In patients with unilateral BM, ipsilateral and contralateral CP volumes were compared by paired t-test and Wilcoxon signed-rank test (paired Cohen’s dz reported); the proportion with ipsilateral predominance was tested against 0.5 by exact binomial test. A signed CPAI [(L−R)/(L+R)] was compared across laterality subgroups (left-only, right-only, bilateral, midline) by Kruskal–Wallis test, with one-sample Wilcoxon tests against zero within each subgroup, multiplicity across the four subgroup tests being controlled by Benjamini–Hochberg FDR. |CPAI| magnitude was compared between each laterality subgroup and non-BM controls by Mann–Whitney U test. Additionally, logistic regression tested whether signed CPAI predicted right-only versus left-only metastasis. ***(iv) Histological grade stratification.*** To assess whether CPAI is independent of primary-tumour aggressiveness, the following were evaluated in patients with available grade (n = 671, 67.1%): the CPAI–grade association (Pearson correlation); grade as an independent covariate in a multivariable model adjusting for CPAI and CA15-3; and grade-stratified CPAI ORs, compared via a CPAI × grade product-term interaction test.

**Single-marker decomposition and effect-modification analyses.** Two pre-specified secondary analyses assessed whether the CPAI–BM association is concentrated in or modified by individual peripheral markers. Single-marker decomposition used product-of-coefficients estimation with 5,000 bias-corrected and accelerated (BCa) bootstrap confidence intervals and Sobel tests to quantify the share of the CPAI–BM association attributable to each of the 10 markers. Stratified CPAI × marker interaction analyses dichotomised each marker at its cohort-specific median; within-stratum CPAI ORs were estimated by logistic regression, and interaction P-values were derived from the corresponding product terms. Multiplicity across both analyses was controlled by Benjamini–Hochberg false discovery rate (FDR) at q < 0.05 over the 10 markers in each family.

### Supplementary Figure

#### Figure S1. Visual Comparison of Choroid Plexus Segmentation Performance Across Different Deep Learning Models

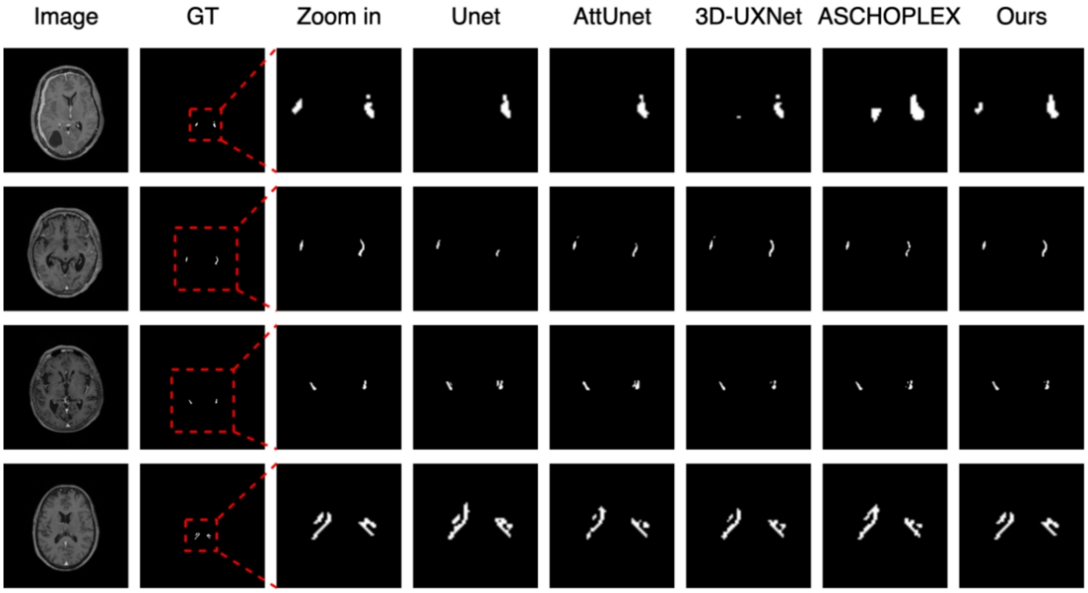


Representative axial slices from four different subjects demonstrating choroid plexus segmentation results. Columns from left to right: Image, original T1-weighted contrast-enhanced MRI; GT, ground truth manual segmentation; Zoom in, magnified view; subsequent columns show segmentation outputs from Unet, AttUnet, 3D-UXNet, ASCHOPLEX, and Ours. White regions represent segmented choroid plexus.

### Supplementary Tables

#### Table S1. Univariate logistic regression for brain metastasis.

| Predictor (per +1 SD) | OR | 95% CI | P |
|---|---|---|---|
| CPAI | 1.53 | 1.33–1.77 | $5.0 \times 10^{-9}$ |
| Age | 0.94 | 0.80–1.10 | 0.415 |
| log(diagnostic-to-MRI interval) | 3.40 | 2.75–4.19 | $4.0 \times 10^{-30}$ |
| NLR | 1.42 | 1.22–1.66 | $8.5 \times 10^{-6}$ |
| MLR | 1.68 | 1.43–1.97 | $1.8 \times 10^{-10}$ |
| PLR | 1.18 | 1.00–1.38 | 0.045 |
| SII | 1.08 | 0.92–1.27 | 0.326 |
| SIRI | 1.37 | 1.17–1.60 | $8.0 \times 10^{-5}$ |
| PIV | 1.13 | 0.96–1.32 | 0.142 |
| NHR | 1.09 | 0.93–1.28 | 0.269 |
| LHR | 0.70 | 0.60–0.82 | $9.4 \times 10^{-6}$ |
| MHR | 1.26 | 1.07–1.47 | 0.005 |
| PHR | 0.82 | 0.70–0.96 | 0.012 |
| CEA | 1.83 | 1.57–2.13 | $4.1 \times 10^{-15}$ |
| CA15-3 | 1.93 | 1.65–2.24 | $2.5 \times 10^{-17}$ |

Each row is a separate logistic regression of brain metastasis status on the single predictor named. Predictors were log-transformed where skewed and standardised on the full cohort, so every odds ratio is per +1 SD. OR, odds ratio; CI, confidence interval. Marker abbreviations as in Table 1.

#### Table S2. Cross-correlation between CPAI and the ten peripheral markers.

| Marker | Axis | Pearson r with CPAI | P |
|---|---|---|---|
| NLR | Inflammation | +0.030 | 0.337 |
| MLR | Inflammation | +0.038 | 0.226 |
| PLR | Inflammation | +0.007 | 0.817 |
| SII | Inflammation | +0.015 | 0.632 |
| SIRI | Inflammation | +0.009 | 0.785 |
| PIV | Inflammation | −0.001 | 0.968 |
| NHR | Oxidative stress | +0.019 | 0.543 |
| LHR | Oxidative stress | −0.008 | 0.800 |
| MHR | Oxidative stress | +0.014 | 0.656 |
| PHR | Oxidative stress | +0.005 | 0.884 |

No correlation exceeds 0.05 in absolute value; the largest is +0.038 for MLR. The complete pairwise matrix among CPAI and the ten markers, including the high correlations among the markers themselves, is displayed in Figure 2B.

### Table S3. Collinearity diagnostics for the primary multivariable model.

| Variable | VIF | Interpretation |
|---|---|---|
| CPAI | 1.02 | Acceptable |
| log(diagnostic-to-MRI interval) | 1.10 | Acceptable |
| Age | 1.09 | Acceptable |
| CA15-3 | 1.43 | Acceptable |
| CEA | 1.45 | Acceptable |
| MLR | 1.26 | Acceptable |
| LHR | 1.20 | Acceptable |

MLR and LHR were retained as the representative markers of the inflammatory–monocyte and lipid–oxidative-stress axes because 9 of the 45 pairwise correlations among the ten markers exceeded 0.70 in absolute value, which precluded entering all ten. Every variance inflation factor in the fitted model falls below 1.50, so the retained set introduces no material collinearity. VIF, variance inflation factor.

### Table S4. Sensitivity analyses of the CPAI–brain metastasis association.

| Model and cohort | Adjusted OR | 95% CI | P |
|---|---|---|---|
| Primary model, full cohort<br>CPAI + age + log(dx interval) + MLR + LHR + CEA + CA15-3 | 1.58 | 1.32–1.88 | $3.6 \times 10^{-7}$ |
| Parsimonious model, full cohort<br>CPAI + age + log(dx interval) + CEA + CA15-3 | 1.60 | 1.34–1.90 | $1.4 \times 10^{-7}$ |
| Extended model, full cohort<br>primary model + histological grade | 1.63 | 1.31–2.03 | $9.2 \times 10^{-6}$ |
| Propensity-score matched, caliper 0.15 | 1.73 | 1.36–2.19 | $7.8 \times 10^{-6}$ |
| Propensity-score matched, caliper 0.20 | 1.73 | 1.36–2.19 | $7.8 \times 10^{-6}$ |
| Propensity-score matched, caliper 0.25 (primary) | 1.73 | 1.36–2.19 | $7.8 \times 10^{-6}$ |
| Propensity-score matched, caliper 0.30 | 1.73 | 1.36–2.19 | $7.8 \times 10^{-6}$ |

All odds ratios are for CPAI per +1 full-cohort SD. Propensity scores were estimated from age and log(diagnostic-to-MRI interval) by logistic regression, with 1:1 optimal matching without replacement that globally minimises the total within-pair distance in the logit propensity score; balance was confirmed at a standardised mean difference below 0.10 for both covariates. The four calipers between 0.15 and 0.30 selected the same matched set, so their estimates coincide. dx interval, diagnostic-to-MRI interval.

### Table S5. Choroid plexus volume and asymmetry by brain metastasis laterality.

| Group | n | Left CP (mm³) | Right CP (mm³) | Signed CPAI, mean (95% CI) | \|CPAI\|, median (IQR) |
|---|---|---|---|---|---|
| Control, no BM | 810 | 523.3 ± 176.0 | 520.4 ± 166.2 | 0.000 (−0.008, +0.007) | 0.059 (0.028–0.114) |
| Left-only BM | 25 | 531.1 ± 217.1 | 513.1 ± 192.7 | +0.005 (−0.051, +0.062) | 0.071 (0.012–0.119) |
| Right-only BM | 14 | 560.6 ± 230.9 | 504.3 ± 224.5 | +0.058 (−0.016, +0.133) | 0.064 (0.040–0.163) |
| Bilateral BM | 141 | 504.2 ± 212.9 | 485.4 ± 199.2 | +0.010 (−0.019, +0.039) | 0.103 (0.043–0.176) |
| Midline BM | 10 | 433.1 ± 165.1 | 490.6 ± 179.6 | −0.092 (−0.193, +0.010) | 0.061 (0.045–0.091) |

Volumes are mean ± SD. Signed CPAI is (left − right) / (left + right), so a positive value indicates a larger left plexus. The unilateral subgroup used in the paired analysis is the union of the left-only and right-only rows. In that subgroup the ipsilateral and contralateral volumes did not differ (paired t-test P = 0.66; Wilcoxon signed-rank P = 0.62; paired Cohen's dz = −0.07), and the proportion with ipsilateral predominance was indistinguishable from chance (exact binomial P = 1.00). Signed CPAI did not differ across the four brain metastasis subgroups (Kruskal–Wallis P = 0.11) and no subgroup differed from zero after Benjamini–Hochberg correction across the four one-sample Wilcoxon tests (all q ≥ 0.20). Against controls, |CPAI| was elevated in bilateral brain metastasis (Mann–Whitney P < 0.001) but not in unilateral brain metastasis (pooled P = 0.52; left-only versus control P = 0.94, right-only versus control P = 0.23). Logistic regression using signed CPAI to discriminate right-only from left-only metastasis was not significant (P = 0.28). BM, brain metastasis; CP, choroid plexus; IQR, interquartile range.

### Table S6. CPAI–brain metastasis association stratified by histological grade.

| Grade stratum | n | BM prevalence | CPAI OR (95% CI) | P |
|---|---|---|---|---|
| Grade I, well differentiated | 27 | 3.7% | Not estimable | — |
| Grade II, moderately differentiated | 461 | 14.1% | 1.46 (1.19–1.79) | <0.001 |
| Grade III, poorly differentiated | 183 | 19.7% | 1.91 (1.23–2.97) | 0.004 |
| Grade not available | 329 | 26.7% | 1.61 (1.23–2.11) | <0.001 |
| Overall cohort | 1,000 | 19.0% | 1.53 (1.33–1.77) | <0.001 |

Odds ratios are for CPAI per +1 full-cohort SD, estimated within each stratum. Grade I is not estimable because the stratum contains a single brain metastasis case. Within the graded patients CPAI was uncorrelated with grade (Pearson r = 0.004, P = 0.93); grade entered as an independent covariate alongside CPAI and CA15-3 remained associated with brain metastasis (per-grade OR 1.84, 95% CI 1.18–2.85, P = 0.007); and the CPAI × grade product term was not significant (P = 0.33). The corresponding extended multivariable model appears in Table S4.

### Table S7. Single-marker decomposition of the CPAI–brain metastasis association.

| Marker | Axis | Path a | Path b | Indirect | Share (%) | Sobel P | BCa 95% CI | BCa P | q |
|---|---|---|---|---|---|---|---|---|---|
| NLR | Inflammation | 0.059 | 0.338 | 0.020 | 4.6 | 0.090 | 0.001 to 0.049 | 0.063 | 0.217 |
| MLR | Inflammation | 0.060 | 0.512 | 0.031 | 7.2 | 0.069 | 0.001 to 0.071 | 0.065 | 0.217 |
| PLR | Inflammation | 0.004 | 0.168 | 0.001 | 0.2 | 0.889 | −0.011 to 0.018 | 0.892 | 0.892 |
| SII | Inflammation | 0.032 | 0.067 | 0.002 | 0.5 | 0.529 | −0.003 to 0.020 | 0.664 | 0.892 |
| SIRI | Inflammation | 0.064 | 0.295 | 0.019 | 4.4 | 0.078 | 0.002 to 0.047 | 0.046 | 0.217 |
| PIV | Inflammation | 0.044 | 0.099 | 0.004 | 1.0 | 0.364 | −0.003 to 0.023 | 0.417 | 0.834 |
| NHR | Oxidative | 0.050 | 0.068 | 0.003 | 0.8 | 0.466 | −0.005 to 0.022 | 0.577 | 0.892 |
| LHR | Oxidative | −0.012 | −0.366 | 0.004 | 1.0 | 0.705 | −0.021 to 0.035 | 0.752 | 0.892 |
| MHR | Oxidative | 0.049 | 0.213 | 0.010 | 2.4 | 0.185 | 0.000 to 0.032 | 0.108 | 0.270 |
| PHR | Oxidative | −0.008 | −0.199 | 0.002 | 0.4 | 0.807 | −0.013 to 0.021 | 0.822 | 0.892 |

Product-of-coefficients decomposition of the total CPAI effect on brain metastasis, which was 0.428 on the log-odds scale. Path a is the CPAI-to-marker coefficient and path b the marker-to-outcome coefficient given CPAI; the indirect term is their product and the share column expresses it as a percentage of the total effect. BCa confidence intervals and BCa P values are from 5,000 bias-corrected and accelerated bootstrap resamples; Sobel P values are from the normal-approximation test; q is the Benjamini–Hochberg adjusted BCa P across the ten markers. No marker reaches q < 0.05 under either the Sobel or the bootstrap criterion. Oxidative, oxidative stress; BCa, bias-corrected and accelerated.

### Table S8. Stratified CPAI × marker interaction analyses.

| Marker | Axis | CPAI OR (95% CI), low stratum | CPAI OR (95% CI), high stratum | P for interaction | q |
|---|---|---|---|---|---|
| NLR | Inflammation | 1.28 (0.98–1.67) | 1.82 (1.44–2.29) | 0.054 | 0.136 |
| MLR | Inflammation | 1.18 (0.88–1.59) | 1.81 (1.45–2.26) | 0.037 | 0.123 |
| PLR | Inflammation | 1.34 (1.06–1.69) | 1.93 (1.49–2.50) | 0.034 | 0.123 |
| SII | Inflammation | 1.55 (1.21–1.99) | 1.61 (1.28–2.02) | 0.778 | 0.975 |
| SIRI | Inflammation | 1.42 (1.09–1.85) | 1.67 (1.34–2.09) | 0.462 | 0.771 |
| PIV | Inflammation | 1.51 (1.18–1.92) | 1.62 (1.29–2.04) | 0.789 | 0.975 |
| NHR | Oxidative stress | 1.71 (1.32–2.20) | 1.48 (1.19–1.84) | 0.443 | 0.771 |
| LHR | Oxidative stress | 1.96 (1.53–2.51) | 1.22 (0.94–1.57) | 0.012 | 0.119 |
| MHR | Oxidative stress | 1.56 (1.18–2.06) | 1.57 (1.27–1.93) | 0.975 | 0.975 |
| PHR | Oxidative stress | 1.60 (1.27–2.03) | 1.56 (1.23–1.98) | 0.911 | 0.975 |

Each marker was dichotomised at its cohort median and the CPAI odds ratio estimated within each stratum, adjusted for log(diagnostic-to-MRI interval). Interaction P values come from the corresponding CPAI × marker product term; q is the Benjamini–Hochberg adjusted value across the ten markers. No interaction survives adjustment. The smallest adjusted value, 0.119, is for LHR, which also has the smallest unadjusted P.

**Table S9. Performance Comparison of Choroid Plexus Segmentation Models Using Five-Fold Cross-Validation**

| Model | Dice (↑) | P | HD95 (↓) | P | Recall (↑) | P | Precision (↑) | P |
|---|---|---|---|---|---|---|---|---|
| Unet | 0.803±0.039 | <0.001 | 2.878±1.283 | <0.001 | 0.769±0.060 | <0.001 | 0.871±0.010 | <0.001 |
| AttUnet | 0.816±0.035 | <0.001 | 2.315±1.019 | <0.001 | 0.818±0.043 | 0.002 | 0.839±0.013 | <0.001 |
| 3D-UXNet | 0.818±0.035 | <0.001 | 2.548±1.112 | <0.001 | 0.799±0.058 | <0.001 | 0.844±0.043 | <0.001 |
| ASCHOPLEX | 0.815±0.044 | <0.001 | 2.331±1.172 | <0.001 | 0.813±0.069 | <0.001 | 0.853±0.058 | 0.002 |
| **Ours** | **0.830±0.030** | — | **1.902±0.677** | — | **0.829±0.053** | — | **0.882±0.026** | — |

Dice, Dice similarity coefficient; HD95, 95th percentile Hausdorff distance in mm. Bold row indicates the chosen model ("Ours"). All P-values correspond to paired comparisons against the chosen model.
